# *Clostridioides difficile* MreE (PBP2) variants facilitate clinical disease during cephalosporin exposures

**DOI:** 10.1101/2023.10.23.23297415

**Authors:** Jay Noboru Worley, Nicholas D. Benedetto, Mary Delaney, Ana Oliveira Paiva, Marie-Pierre Chapot-Chartier, Johann Peltier, Lynn Bry

## Abstract

Cephalosporins are the most common triggers of healthcare-associated *Clostridioides difficile* infections (CDI). Here, we confirm gene-level drivers of cephalosporin resistance and their roles in promoting disease. Genomic-epidemiologic analyses of 306 *C. difficile* isolates from a hospital surveillance program monitoring asymptomatic carriers and CDI patients identified prevalent third-generation cephalosporin resistance to ceftriaxone at >256 ug/mL in 26% of isolates. Resistance was associated with patient cephalosporin exposures 8-10 days before *C. difficile* detection. Genomic analyses identified variants in the *mreE* penicillin binding protein 2 (PBP2) associated with resistance to multiple beta-lactam classes. Transfer of variants into susceptible strain CD630 elevated resistance to first and third-generation cephalosporins. Transfer into the mouse-infective strain ATCC 43255 enabled disease when mice were exposed to 500ug/mL cefoperazone, a dose that inhibited the isogenic susceptible strain. Our findings establish roles of cephalosporins and *mreE*-cephalosporin-resistant variants in CDI and provide testable genetic loci for detecting resistance in patient strains.

## Introduction

Antibiotic exposures enhance risks for healthcare-associated *Clostridioides difficile* infections (CDI), particularly for patients who receive lincosamides, fluoroquinolones, or cephalosporins in the course of their clinical care, antibiotics commonly prescribed for other conditions [1–3]. *C. difficile* is often multidrug resistant (MDR) from acquired genes such as *ermB* for clindamycin and macrolide resistance, and from chromosomal mutations such as ones in the *gyrA* DNA gyrase that mediate resisstance to fluoroquinolones. However, the role of *C. difficile* resistance in its colonization and progression to disease in patients with antibiotic-depleted microbiota, has remained ill-defined, but remains a critical cquestion to understand how patient antibiotic exposures and strain-level resistance impact patient risks for *C. difficile* colonization and disease [4].

Multiple gene-level factors have been evaluated as drivers in *C. difficile’s* cephalosporin resistance. An intrinsic class D beta-lactamase providing degress of resistance across genomically diverse *C. difficile* has been identified [5, 6]. Variants in the pathogen’s penicillin-binding protein 2 (PBP2) have also been identified that have reduced binding-affinity to certain beta-lactam classes, including cephalosporins and carbapenems [7]. However, causative effects of these variants on phenotypic resistance *in vitro* and *in vivo* have not yet been defined, limiting our understanding of their role in patient disease [8–10].

To identify contributions of genotypes on phenotypic antibiotic restance and on patient colonization and disease, we evaluated 306 clinical *C. difficile* isolates from asymptomatic carriers and actively infected patients [11] for antibiotic resistance to disease-triggering and therapeutic antibiotics. Ceftriaxone resistance at ≥256 µg/ml was associated with patients who had received cephalosporins as part of their clinical care 8-10 days prior to initial detection of *C. difficile* colonization or symptomatic disease. Genomic analyses identified amino acid substitutions in the *mreE* gene, a member of the *mre* operon that encodes a PBP2. Transfer of *mreE* resistant alleles *in trans* to the susceptible type strains CD630 and ATCC 43255 confirmed effects of *mreE* variants on phenotypic resistance to different beta-lactam antibiotics. *In vivo* studies of a highly resistant *mreE* variant transferred into the mouse-infective strain ATCC 43255 demonstrated the ability of *mreE* mediated cephalosporin resistance to facilitate CDI in mice receiving the third-generation cephalosporin cefoperazone. Our findings confirm contributions of *mreE* variants in cephalosporin resistance and their roles *in vivo* to facilitate *C. difficile* colonization and disease.

## Methods

### Clinical isolate and data collection

Isolates in this study were collected under Institutional Review Board protocol 2011-P-002883 (L. B., Partners Healthcare) and described in Worley, et al. [11]. Patient data was collected from the Mass General Brigham Research Patient Data Registry and deidentified for analyses [12]. Patient admission status and antibiotic therapy were analyzed per calendar day. For risks of antibiotic therapy on *C. difficile* acquisition, as defined by the point at which patients were found to be asymptomatically colonizated or infected with toxigenic *C. difficile*, a five-day window centered on each day was evaluated for all patients receiving a given antibiotic within the window.

### Antimicrobial resistance tests

Antimicrobial resistance testing used the methods in the Clinical and Laboratory Standards Institute (CLSI) [13]. ETESTs (bioMérieux, Durham, NC) were performed on Brucella agar with 5% sheep blood, hemin, and vitamin K (Remel, Lenexa, KS). Broth microdilution experiments were performed in at least 2 independent replicates in TY broth at pH 7.4 containing 30 g/L tryptose (MilliporeSigma, Burlington, MA), 20 g/l yeast extract (Thermo Fisher Scientific, Waltham, MA), and 0.1 g/l sodium thioglycolate (Sigma-Aldrich, Burlington, MA). Antibiotics were dissolved directly into TY media at the highest dose, filter sterilized, then serially diluted in TY media to prepare the broth dilution series, using the following antibiotics: ceftriaxone sodium salt hemi(heptahydrate) cefotaxime sodium salt, cefoxitin sodium salt, cefazolin sodium salt (Acros Organics, Waltham, MA), amoxicillin sodium salt, cefepime, and cefpodoxime (Alfa Aesar, Ward Hill, MA), cephalexin (ApexBio Technology, Houston, TX), ampicillin (Sigma-Aldrich, Burlington, MA), USP grade cefoperazone sodium salt (MP Biomedicals, Santa Ana, CA), and meropenem (Ark Pharm, Libertyville, IL).

### Phylogenetic analyses

Whole-genome single nucleotide polymorphisms (SNPs) were identified as described in Worley, et al. [11]. Identification of strain gene homologs of target genes used the ATCC 43255 reference genome, NCBI RefSeq NZ_CP049958 [14]. Antimicrobial resistance genes were identified using AMRFinder [15]. PATRIC annotation and literature review identified a set of 88 peptidoglycan-associated encoded protein sequences [16]. tBLASTn from BLAST 2.7.1+ identified homologous genes among the genomes [17]. MUSCLE 3.8.1551 generated sequence alignments [18], and FastTree 2.1.3 performed phylogenetic analyses of encoded protein sequences or SNP matrices [19]. Phylogenetic trees were visualized using iTOL [20]. To identify protein sequence variants associated with enhanced resistance, 88 known cell wall or peptidoglycan-associated proteins were evaluated among strains using Fisher’s exact test (Table S2).

### Reverse-transcriptase PCR

Table S1 details the primers and probes used for reverse-transcriptase PCR to detect *in vitro* gene expression of *mreE.* Acetate kinase was used as an expression control. Microbial RNA was harvested from flash-frozen pellets of log-phase TY broth liquid after anaerobic growth for four hours, and using the Zymo Direct-zol RNA purification kit (Zymo, Irvine, CA). RtPCR reactions used SuperScript II Reverse Transcriptase (Invitrogen, Waltham, MA). PCR for the amplification of the *mreD*-*mreE* junction sequence and relative expression levels by SYBR used the PowerUp SYBR Green Master Mix (Applied Biosystems, Waltham, MA). Probe displacement assays by TaqMan used TaqMan Universal Master Mix II (Applied Biosystems, Waltham, MA). Probes for the *mreE* V497 and L497 alleles used a 5’ 6-FAM or SUN fluorophore, respectively, with a 3’ Iowa Black FQ quencher (Integrated DNA Technologies, Coralville, IA) and corresponding PCR primers (Integrated DNA Technologies, Coralville, IA) (Table S1). PCR reactions used a QuantStudio 12K Flex (Applied Biosystems, Scientific, Waltham, MA).

### Purification and structural analysis of peptidoglycan

Peptidoglycan was extracted from *C. difficile* strains as described in Peltier *et al.* with the following modifications [21]. *C. difficile* cultures grown to OD_600_≈1 at 37°C in tryptone yeast extract medium (TY) were chilled on ice and cells were harvested by centrifugation. Cell pellets were resuspended in cold H_2_O, boiled for 10min, cooled again, and centrifuged. The cells were boiled in 4% sodium dodecyl sulfate, washed several times with H_2_O, and processed in a FastPrep apparatus (MP Bioscience) for 30 s at 4 m/s to disrupt the cells. The insoluble material was sequentially treated with pronase, α–amylase, DNase, RNase and trypsin to purify the cell wall and then incubated with 48% hydrofluoric acid at 4 °C for 16 h to remove wall polysaccharides. Purified PG was digested with mutanolysin (Sigma), and the soluble muropeptides were reduced with sodium borohydride. Peptides were then separated by reverse phase-ultra high-pressure liquid chromatography (RP-UHPLC) with a 1290 chromatography system (Agilent Technologies, Santa Clara, CA, United States) and a ZORBAX Eclipse Plus C18 RRHD column (100 by 2.1mm; particle size, and 1.8μm; Agilent Technologies) at 50°C using ammonium phosphate buffer and methanol linear gradient to evluate the peptidoglycan fractions.

### *mre* operon cloning and conjugation into recipient strains

*mre* operon sequences including the *mre* genes and native 5’ promoter sequences were amplified using the primers specified in Table S1 (Integrated DNA Technologies, Coralville, IA) and amplified using Platinum SuperFi II (Invitrogen, Waltham, MA) with a 54 °C annealing temperature for 10 cycles and a 57 °C annealing temperature for 30 cycles. All cycles used a 4 minute extension time at 72°C. PCR cleanup was performed using the QIAquick Gel Extraction Kit on a 0.8% agarose gel (Qiagen, Hilden, Germany). Insertions were made via XbaI and KpnI digestion into pMTL84151 using Ligase Version T4 DNA Ligase (New England Biolabs, Ipswich, MA), with orientation towards the plasmid’s transcriptional terminator [22]. Ligation products were transformed into NEB5a cells (New England Biolabs, Ipswich, MA) and evaluated for blue or white colony color on LB agar (BD Biosciences, Franklin Lakes, NJ) supplemented with IPTG (Sigma Chemical, St. Louis, MO) and X-gal (Invitrogen, Waltham, MA). Plasmid purification used the QIAprep Spin Miniprep Kit (Qiagen, Hilden, Germany). Resulting plasmids were confirmed by sequencing and transformed by electroporation into *E. coli* S17-1 [23][24]. Transformation into *C. difficile* CD630 and ATCC 43255 was performed by suspending 48 hour *C. difficile* cultures on TY solid media and overnight S17-1 on LB solid media with chloramphenicol into reduced TY liquid media, without antibiotics, to approximately OD_600_ 0.1. The cultures were then mixed and 100 µl spotted onto reduced TY media and incubated overnight anaerobically at 37°C. Growth from the mating reactions was scraped and resuspended into 1 ml reduced TY, then 100µl plated onto TY with chloramphenicol and oxacillin.

To evaluate whether the constructs caused overexpression of *mreE*, we evaluated the expression of the chromosomal and plasmid-borne alleles in the CD630 and ATCC 43255 transconjugants, relative to the acetate kinase housekeeping gene *ackA* (Fig S1). Among strains, expression of the plasmid-borne *mre* locus comprised approximately half of *mreE* expression (Fig S2), indicating that expression *in trans* impacted phenotypic resistance and associated *in vivo* phenotypes. Further, the total expression of *mreE* was not significantly altered from the parent or recipient strains.

### Mouse experiments

All mouse experiments were conducted under an institutional IACUC protocol. Conventionally colonized 5-week old male Swiss Webster mice (Taconic Biosciences, Germantown, NY) were singly housed throughout the experiments. Antibiotic treatments consisted of USP grade cefoperazone at, at 96, 250 or 500ug/mL, to mirror human doses and dissolved in water containing 3% sucralose (Alfa Aesar, Ward Hill, MA) to mask any bitter tastes. All mice were provided ad libitum access to the antibiotic drinking water, or control 3% sucralose in water, from five days prior to 14 days after challenge. Control mice for symptomatic infections received a single 10 mg/kg intraperitoneal injection of USP grade clindamycin (Sigma Chemical, St. Louis, MO) given 24h before challenge, and control 3% sucralose drinking water thereafter. Mice were gavaged with 10^3^ CFU *C. difficile* spores [14]. Mice were weighed and scored for body condition at the same time on days −5, −1, 0, 1, 2, 3, 5, 7, 10, and 14. Fecal pellets were collected on days −5, 0, 1, 2, 3, 4, and 5 during acute infection.

### 16S rRNA phylotyping analyses

Fecal DNA was extracted using the Zymo Quick-DNA Fecal/Soil Microbe Miniprep Kit (Zymo, Irvine, CA).16S rRNA gene sequencing of the v4 region was performed by Illumina MiSeq in the Massachusetts Host Microbiome Center. Analyses used DADA2 1.14.1 with the Silva nr99 v138.1 training set, additionally using Phyloseq 1.30.0 and Biostrings 2.54.0 to classify identified OTUs [25–29]. Analyses of diversity were performed in Python 3.7.6 using Scikit-bio 0.5.6 [30].

### Toxin B enzyme-linked immunosorbent assay (ELISA)

ELISAs for toxin B levels in fecal pellets were conducted as previously described with the following modifications [14]. Fecal pellets were homogenized into 1ml sterile PBS and toxin levels normalized to pellet mass.

### Statistical tests and visualization

Antibiotic resistance histograms and bar charts, mutation maps, 16S rRNA visualization and real-time PCR graphs were created using Matplotlib 3.4.2, and Seaborn [31, 32]. Patient, mouse weight, toxin level data were visualized, and 16S rRNA Shannon diversity index significance tests, were performed using GraphPad Prism version 9.5.1 [33], including these statistical tests: paired t-tests, Welch t-tests, Kruskal-Wallis tests, and controlling FDR by the two-stage step up method of Benjamini, Krieger, and Yekutieli.

## Results

We evaluated 306 unique *C. difficile* isolates from 301 patients monitored in the *C. difficile* surveillance program at Brigham and Women’s Hospital. Strains were evaluated for phenotypic resistance to ceftriaxone, clindamycin, erythromycin, metronidazole, rifampicin, tetracycline, and vancomycin (Fig 1 A-B, Fig S3) [11]. Resistance to the third-generation cephalosporin ceftriaxone occurred most frequently, with 26% of isolates (n=79) demonstrating an MIC of ≥256. µg/ml. While clindamycin resistance at ≥256ug/mL was strongly associated with carriage of the *erm*(B) gene (χ^2^ p < 0.001), phenotypic cephalosporin resistance showed no association with carriage of previously reported beta-lactamase *blaCDD* variants (χ^2^ >0.05) [5, 6, 34].

**Figure 1.**
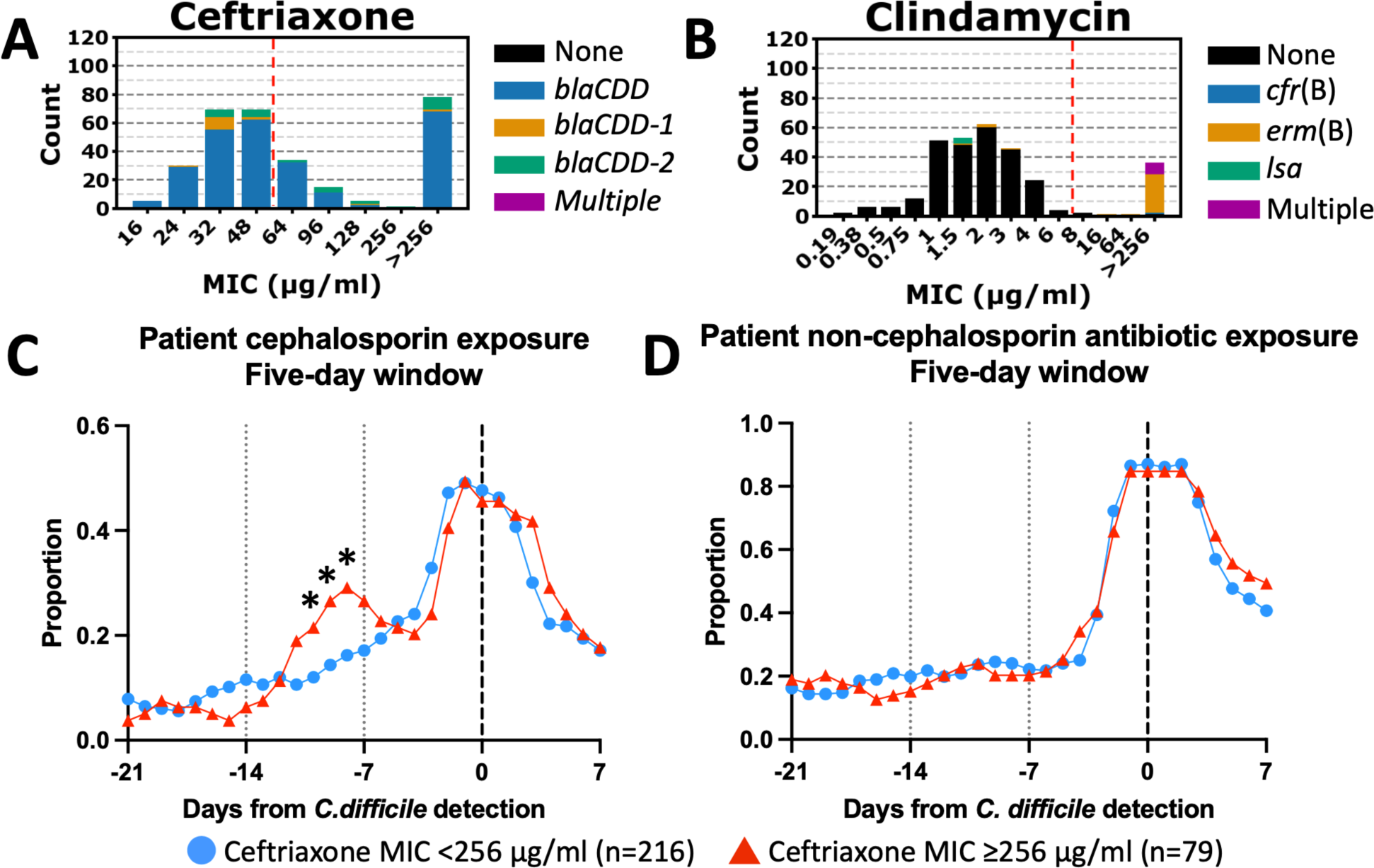
Clinical *C. difficile* isolates with an elevated ceftriaxone MIC are associated with cephalosporin use 8-10 days prior to detection. (A) Number of patient isolates with each ceftriaxone ETEST MIC (n=306). Antibiotic resistance genes from NCBI’s AMR Finder tool at each resistance level are indicated by proportional color in the bar. The CLSI breakpoint is indicated by a dashed red line. (B) The same analysis as in Fig. 1A but assayed for clindamycin ETEST MIC and corresponding antibiotic resistance genes. (C) Proportion of patients receiving a cephalosporin within a five-day window centered on the indicated day (n=295). (D) Proportion of patients receiving a non-cephalosporin antibiotic within a five-day window centered on the indicated day. * indicates a p-value ≤ 0.05 by c^2^ test. For C and D, patients from A and B without complete records were excluded (n=6) as well as secondary isolates from each patient (n=5).

To evaluate contributions of cephalosporin exposures in the acquisition of cephalosporin-resistant *C. difficile*, we evaluated a cohort of 295 patient for whom detailed medical records were available. This cohort included 131 asymptomatically colonized patients (44%) and 164 patients with CDI (55%). Among patient isolates, 79 (27%) had ceftriaxone MICs ≥256 µg/ml. Cephalosporin use eight to ten days prior *to C. difficile* detection was associated with subsequent detection of ceftriaxone-resistant patient isolates (χ^2^ p-values for days 8, 9 and 10 before detection: 0.013, 0.015, 0.040, respectively) (Fig 1C), while patient exposures to non-cephalosporin antibiotics showed no such correlation (Fig 1D). Notably, the occurrence of ceftriaxone-resistant vs. susceptible strains showed no associations with their detection in inpatient or outpatient settings (Fig S4). Among patient beta-lactam exposures prior to *C. difficile* detection, third-generation cephalosporins were the most commonly prescribed (n=93, 32%), followed by fourth-generation (n=60, 20%), first-generation (n=39, 13%), and second-generation cephalosporins (n=3, 1%) (Table 1).

**Table 1.** Cephalosporins prescribed in the 21 days before *C. difficile* detection. Number of patients receiving the stated cephalosporin or generation of cephalosporin in the 21-day period before detection.

| Cephalosporin | Generation | Patients per drug | Patients per generation |
| --- | --- | --- | --- |
| Cefadroxil | First | 3 (1%) | 39 (13%) |
| Cefazolin | First | 36(12%) |  |
| Cephalexin | First | 4 (1%) |  |
| Cefotetan | Second | 2 (1%) | 3 (1%) |
| Cefoxitin | Second | 1 (0%) |  |
| Cefixime | Third | 1 (0%) | 93 (32%) |
| Cefpodoxime | Third | 2 (1%) |  |
| Ceftazidime | Third | 45 (15%) |  |
| Ceftriaxone | Third | 58 (20%) |  |
| Cefepime | Fourth | 60 (20%) | 60 (20%) |

Genomic analyses for beta-lactamases and for amino acid substitutions in genes associated with cell-wall synthesis were investigated for linkages with phenotypic resistance to ceftriaxone. Variants in the MreE protein showed the highest association with phenotypic resistance, and fell within a large genomic clade, designated A, that included as highly resistant sub-clade, designated A1 (Table S2). Clade A contains 47 resistant and three susceptible isolates (odds ratio=114), while sub-clade A1 contains a subset of 25 resistant and 0 susceptible isolates (odds ratio ≥ 108).

The *mreE* gene resides in the *mreBCDE* operon 3’ to *mreD* and separated by an 11bp intergenic region (Fig 2A). Studies by Soutorina, et al. previously identified the start of *mre* operon transcription from a single promoter region upstream of *mreB* [35]. Reverse transcriptase PCR of *mre* gene expression in the susceptible type strain ATCC 43255 and in the resistant clinical isolate CD41 confirmed co-expression of *mreE* with *mreD* and the larger *mre* operon (Fig 2A, Fig S5).

**Figure 2.**
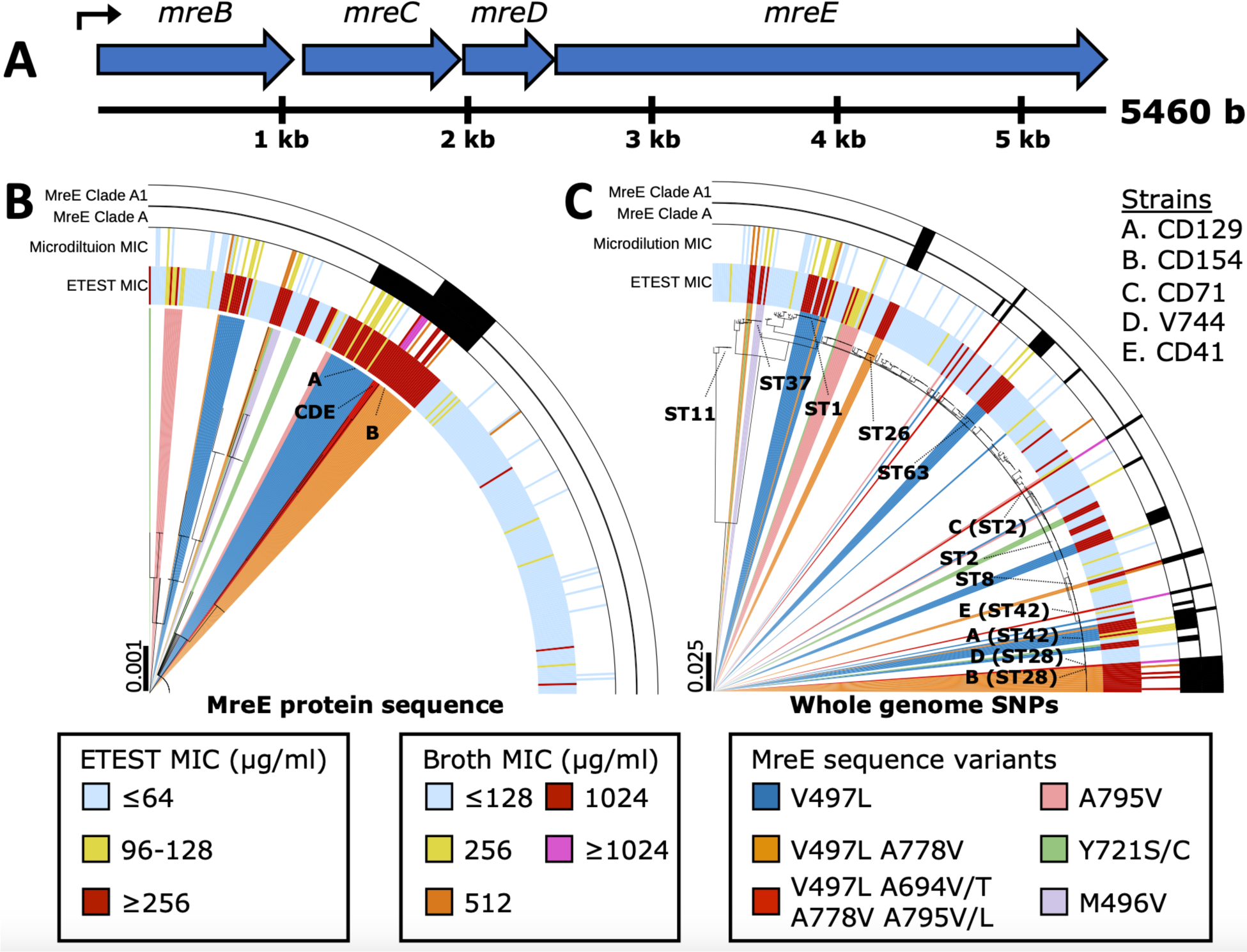
Protein sequence subclades of MreE are linked to ceftriaxone resistance and have convergently evolved. (A) Genetic physical map of the *C. difficile mre* operon protein coding regions. An undefined promoter exists upstream of *mreB*, indicated by a bent arrow. (B) Phylogenetic tree of encoded MreE protein sequences from Fig 1A (n=306). Outer black bars show encoded MreE clades from Table S2. Interior colored bars indicate ceftriaxone MICs as assessed by ETEST and broth microdilution. Colored slices in the dendrogram indicate encoded MreE sequence variants. Specific strains are indicated by letters. (C) Phylogenetic tree of whole-genome SNPs from a core genome alignment for the strains from Fig. 2B. Selected sequence types are indicated in text.

Phylogenetic analysis of MreE protein sequences identified a consistent substitution, V497L, among Clade A strains. In addition, strains within sub-clade A1 carried an additional *mreE* variant, A778V, with V497L (Table S2, Fig 2B). Higher resolution broth microdilution testing of ceftriaxone to 1024 ug/mL showed sub-clade A1 isolates to be at least two times more resistant to ceftriaxone than isolates belonging to Clade A alone.

Among the *C. difficile* whole genome sequence types (wgMLST) obtained from patients, additional clades carried V497L, or V497L and A778V, and demonstrated comparable levels of ceftriaxone resistance, suggesting that independent evolution of the V497L and A778V variants could drive resistance across genomically diverse strains (Fig 2B). Notably, no isolates carried the A778V substitution independently of V497L. Substitutions of Y721 to either serine or cysteine (Y721S/C), and A795 to valine or leucine (A795V/L) were also observed in multiple MreE clades demonstrating phenotypic ceftriaxone resistance. A distinct clade within cephalosporin-resistant strains of ST37 carried the variant M496V adjacent to V497L (Fig 2B, light purple). Lastly, three isolates within sub-clade A1, and demonstrating ceftriaxone resistance ≥1024 µg/ml, harbored additional variants to V497L and A778V: A795V/L, and A694V/T. These findings provide phylogenetic evidence that MreE variants at V497, A778, A795, and Y721 contribute to progressive cephalosporin resistance, with potential contributions from mutations at A694 and M496.

Whole-genome SNP phylogenies confirmed multiple cephalosporin-resistant strain lineages that had independently acquired identical mutations in the MreE protein (Fig 2C). Among *C. difficile* genomic clades demonstrating phenotypic ceftriaxone resistance, 82% of ST1 strains (14 of 17 patient isolates) carried the V497L substitution, with one also carrying A778V. Six of eight ST1 isolates with these mutations demonstrated ceftriaxone resistance ≥256 µg/ml, whereas the two isolates lacking these mutations remained susceptible. Sequence types 42 and 28 were prevalent among cephalosporin resistant isolates, accounting for 21% of all patient isolates (64 of 306 isolates), and 34% of resistant isolates (27 of 79 isolates).

To investigate the effects of *mreE* variants on beta-lactam resistance, we selected representative strains that carried the V497L and additional mutations in *mreE* for functional studies: Clade A isolate CD129 (ST42), and Clade A1 isolates CD154 (ST28), CD71 (ST2), V744 (ST28,) and CD41 (ST42). Genomically, within the *mre* operon, these strains only harbored variants in the *mreE* gene that correlated with increased resistance (Fig 3A). The type strains CD630 and mouse-infective strain ATCC 43255 provided susceptible controls, and recipient strains for gene-transfer studies (Fig 2B, C).

**Fig 3.**
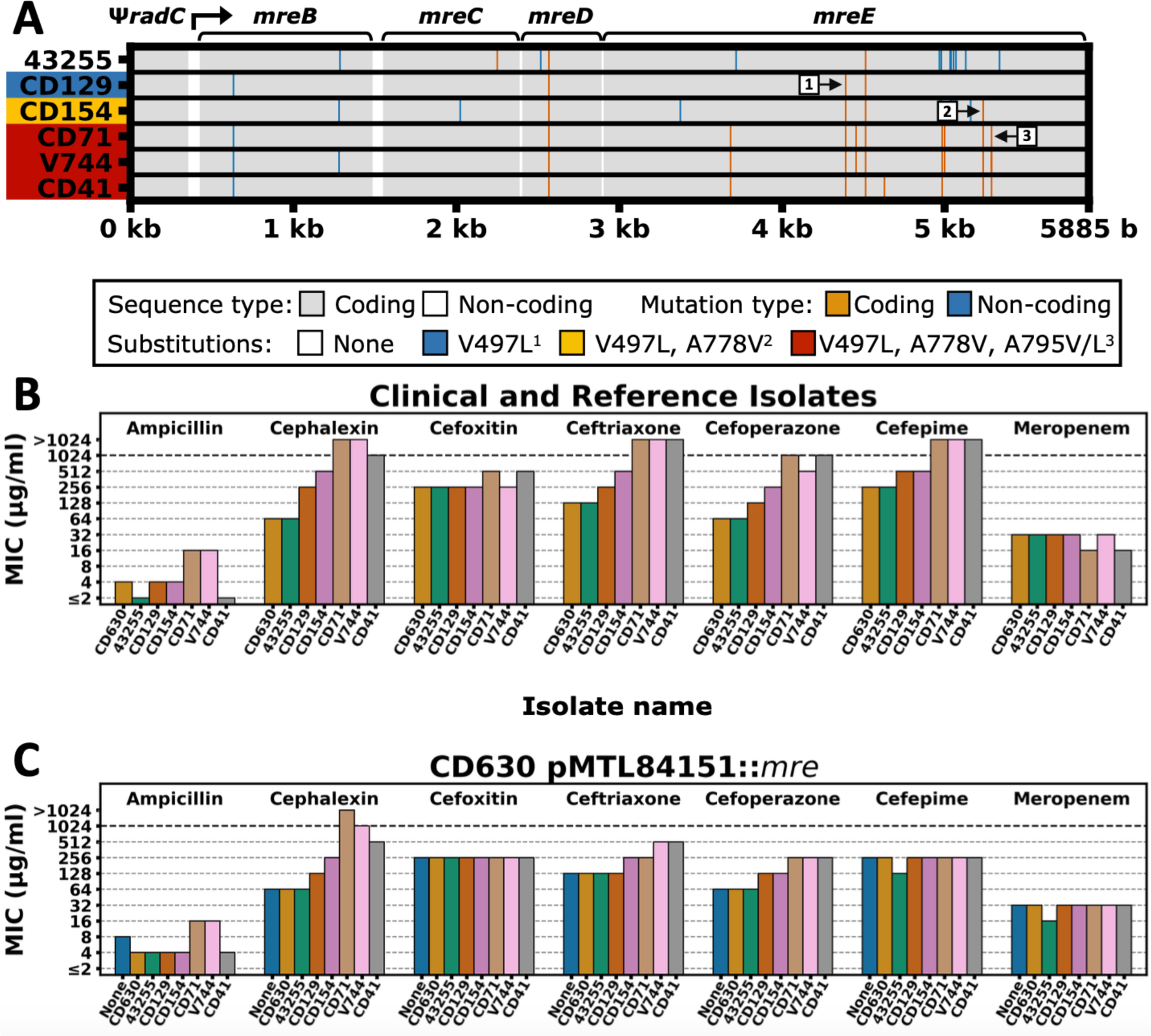
MreE variants show broad resistance to multiple classes of cephalosporins, and phenotypes are partially conferrable by a plasmid-borne *mre* operon. ATCC 43255 is abbreviated 43255. (A) Map of the silent and non-silent mutations in the pMTL84151 insert relative to the ATCC 43255 allele for the indicated strains. Coding regions are indicated by gray blocks, and mutations are represented by vertical lines. The promoter is indicated by a bent arrow. Pointer squares with numbers indicate 1, V497L; 2, A778V; 3, A795V/L. (B) Broth microdilution antibiotic MICs for selected clinical isolates and type strains. (C) Broth dilution antibiotic MICs for *C. difficile* CD630 pMTL84151::*mre* with varying parent strains for the *mre* insert. Except for the empty vector (None), these carry two copies of the *mre* operon, the original chromosomal copy, and another on pMTL84151.

Strains CD71 and V744 demonstrated higher MICs to ampicillin than the other strains, which remained susceptible (Fig 3B). For the first-generation cephalosporin cephalexin, the MIC for CD129 was four-fold that of CD630 and ATCC 43255, CD154 eight-fold, and over eight-fold for CD71, V744, and CD41. In contrast, no consistent increases in resistance were observed for the 2nd generation cephalosporin cefoxitin. For the third-generation cephalosporins ceftriaxone and cefoperazone, CD129 and CD154 demonstrated two and four-fold increases in resistance to both. As with cephalexin, CD71, V744, and CD41 demonstrated ≥eight-fold increases in resistance to third-generation cephalosporins, as well as ≥four-fold increases in resistance to the fourth-generation cephalosporin cefepime. A similar step-wise progression in resistance to cefepime also occurred relative to the accumulation of *mreE* variants. Lastly, no strains demonstrated enhanced resistance to the carbapenem meropenem.

Given MreE’s proposed transpeptidase functions, we analyzed peptidoglycan (PG) structures in the resistant isolates CD154, V744 and CD41, and susceptible strains ATCC 43255 and CD630, to determine if alterations in their PG muropeptides occurred. However, no novel muropeptides, nor consistent changes in their stoichiometry, occurred among susceptible or resistant strains (Fig S6).

For molecular confirmation of the *mre* variants in cephalosporin resistance, we transferred variants *in trans,* into the susceptible and genetically tenable strain CD630. Given *mreE*’s essentiality (CDR20291_0985 in FN545816.1 [36]), and toxicities associated with the over-expression of cell wall synthases, we cloned the full-length 5885 bp *mre* operon, containing the *mre* operon’s promoter region through *mreE* gene (Fig 3A), into pMTL84151 for conjugation into CD630, where the plasmids are denoted by their inserted *mre* operon, such as pCD41 for pMTL84151::*mre*_CD41_ [35, 37].

Strain CD630 carrying plasmid pCD71 or pV744 showed minor increases in ampicillin MICs (Fig 3C). For the first-generation cephalosporin cephalexin, progressive resistance in CD630 transconjugants, and similar to the wild-type strains (Fig 3B), occurred with pCD71, pV744, and pCD41. No increase in resistance occurred for the second generation cephalosporin cefoxitin. For the third-generation cephalosporins ceftriaxone and cefoperazone, CD154, pCD71, pV744, and pCD41 bestowed a 2-4 fold increase in resistance to ceftriaxone, and CD129 did so additionally for cefoperazone. None of the plasmids bestowed increased resistance for the fourth-generation cephalosporin cefepime or the carbapenem meropenem. Thus, *in trans,* the *mreE* variants in pCD71, pV744 and pCD41 showed the greatest impact on increased resistance to ampicillin, the first-generation cephalosporin cephalexin, and third-generation cephalosporins ceftriaxone and cefoperazone.

To evaluate the contributions of host cephlosporin exposures and MreE-mediated cephalosporin resistance on CDI, we first assessed how escalating doses of the third-generation cephalosporin, cefoperazone, depleted the gut microbiota. Cefoperazone was chosen per the availability of a well-validated mouse cefoperazone infection model [38], and per third-generation cephalosporins being the most common antibiotic exposure among *C. difficile* colonized patients in our patient cohort (Table 1).

Exposure to 96, 250 and 500ug/mL of cefoperazone in drinking water showed progressive depletion of the microbiota, in contrast to mice receiving control water (Fig. 4). The cefoperazone-induced shifts also differed from those seen in mice receiving a single intraperitoneal dose of clindamycin, which targeted a broader range of aaerobic taxa [39]. While cefoperazone at 96 ug/mL for 5 days of administration showed no significant effects on the Shannon diversity of the fecal microbiota (p=0.35), progressive reduction in diversity occurred at 250 µg/ml (p=0.029) and 500 µg/ml (p=0.045). Bray-Curtis dissimilarity showed significant differences across cefoperozone doses with progressive variability increasing at 250 µg/ml and 500 µg/ml. Order-level operational taxonomic units (OTU) showed progressive reductions in the relative abundance of taxa in the Bacteroidales and Lachnospirales (Fig 4C) with expansion of taxa associated with the *Clostridia* vadinBB60 group. In contrast, mice exposed to clindamycin demonstrated profound reductions in microbiome diversity, particularly among taxa associated with the Bacteroidales, Lachnospirales and Clostridiales, with increases in relative abundance in the Lactobacillales and Erysipelotrichales (Fig 4C). Per the findings of progressive depletion relative to cefoperazone dosing, we evaluated the course of *C. difficile* disease in mice exposed to 250 versus 500 ug/mL of cefoperazone.

**Figure 4.**
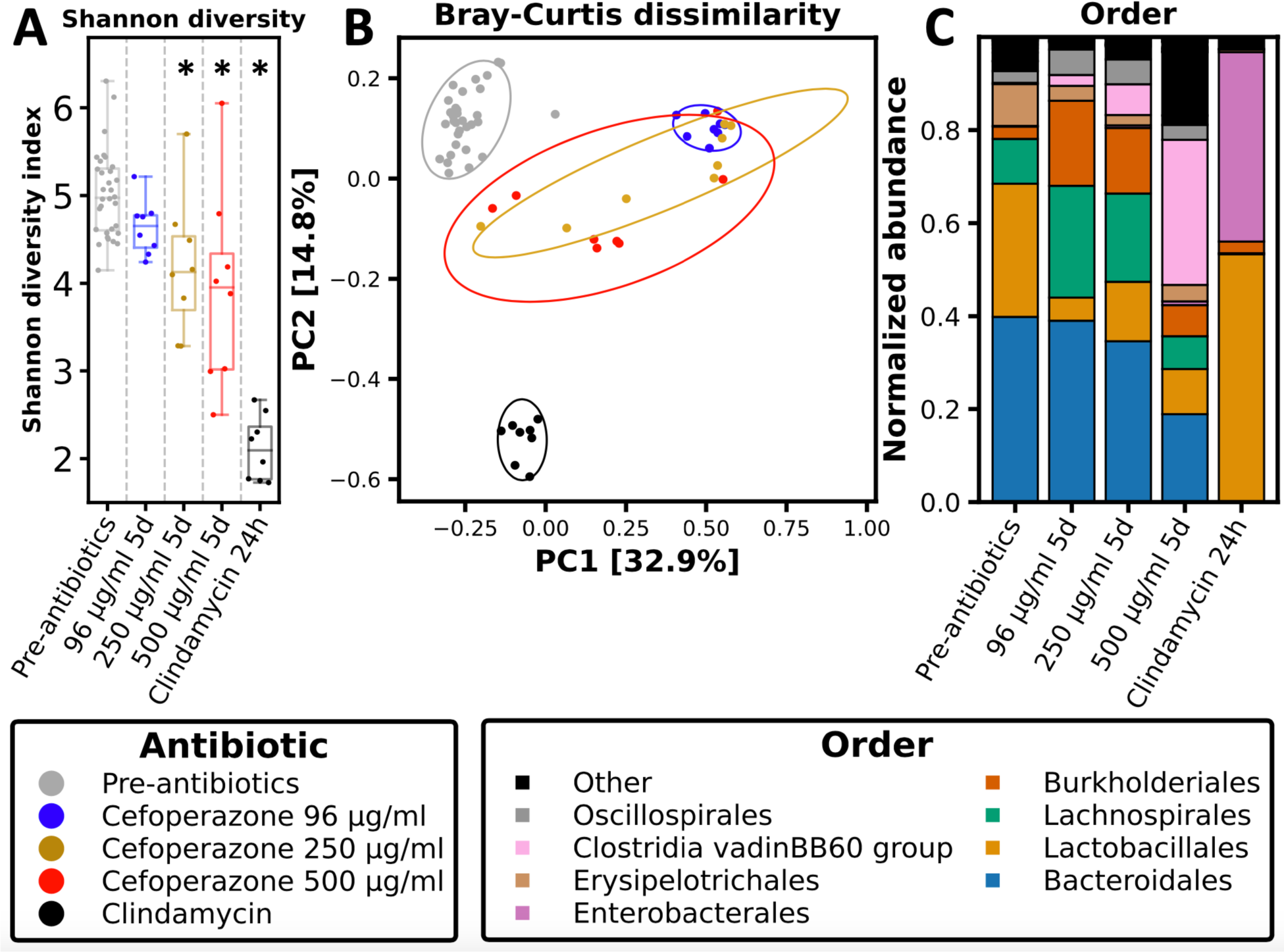
*Ad libitum* cefoperazone in drinking water alters the mouse gut microbiome in a dose dependent manner. The “Pre-antibiotics” state represents fecal samples taken from all mice before antibiotic administration (n=32 mice),.“5d” represents sampling after five days of cefoperazone treatment and before *C. difficile* challenge (n=8 for each group), and “24h” is 24 hours after intraperitoneal injection of clindamycin and prior to *C. difficile* challenge in clindamycin-exposed control mice (n=8). Dose levels in μg/ml refer to cefoperazone. Analyses are based on 16S rRNA sequence data interpreted using DADA2. (A) 16S rRNA Shannon index of diversity. * indicates the post-antibiotic treatment condition is significantly (p ≤ 0.05) different from the pre-antibiotic treatment condition by one-way ANOVA. (B) Principal coordinates analysis of Bray-Curtis dissimilarity. (C) Microbiome composition by phylogenetic Order. * indicates p ≤ 0.05 for a paired t-test.

To evaluate if resistant *mreE* alleles promote infection in an antibiotic-exposed host, we transferred the highly resistant *mre* alleles in pCD41 (Fig 3) to the cephalosporin-susceptible mouse-infective strain ATCC 43255. Transfer of pCD41 into ATCC 43255 bestowed a four-fold increase in ceftriazone MICs compared to ATCC 43255 carrying the control plasmid pMTL84151 or its native *mre* locus in pATCC43255, and a two-fold increase for cefoperazone, similar to the results seen in CD630 (Fig S7). To confirm infectivity of ATCC 43255 harboring pCD41 or pATCC43255, control mice receiving intraperitoneal clindamycin 24h prior to challenge with either strain developed severe infections, confirming that carriage of either plasmid had no effect on strain infectivity (Fig S8).

To mimic clinical antibiotic exposures that enhance risks for *C. difficile* colonization and disease, mice received cefoperazone in drinking water at the selective dose of 500ug/mL, or the non-selective dose at 250 ug/mL, for five days prior to challenge with 1,000 *C. difficile* spores. Animals remained on the given cefoperazone dose after pathogen challenge for the next 14 days (Fig 5A). At the 500 µg/ml cefoperazone dose, mice infected with ATCC 43255, carrying the pCD41 *mre* locus lost significantly more weight than mice infected with ATCC43255 carrying its native *mre* locus in pATCC43255 (Fig 5B). In constrate, no differences occurred between the two strains at the non-selective dose 250 µg/ml where both groups of infected mice showed minimal weight loss (Fig 5C).

**Fig 5.**
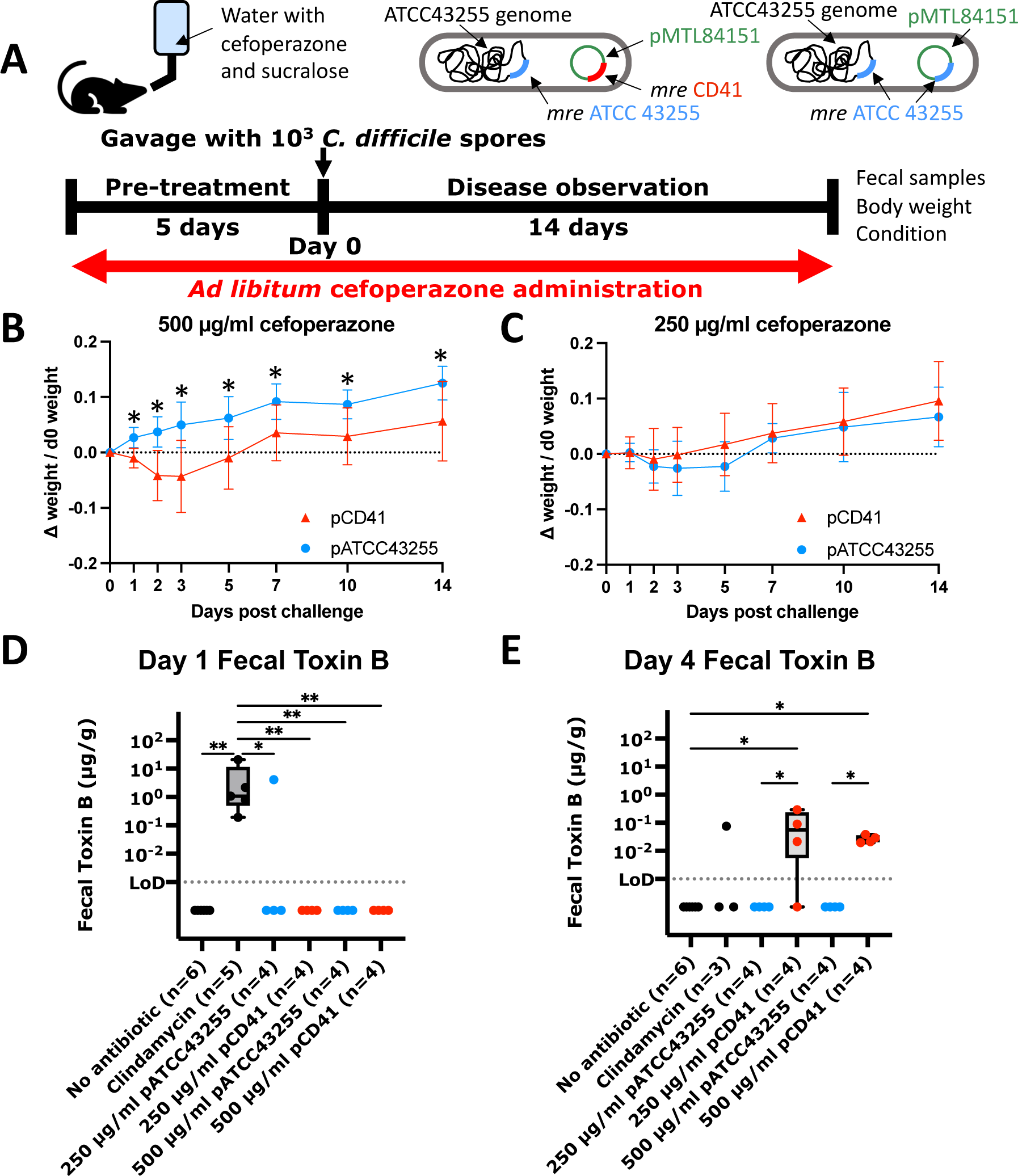
Elevated resistance levels cause CDI. (A) Timeline of cefoperazone experiments. Cefoperazone was administered in drinking water *ad libitum* throughout the timeline. Clindamycin was delivered as a single injection 24h before gavage (not shown). Images representing the mode of antibiotic delivery and a graphical representation of the infecting strains are given above the timeline. Collected data are given to the right. (B-C) Plots of the change in mouse weights between different cohorts, indicated by different colors and symbols (n=8 mice for all cohorts). Error bars are one standard deviation. Values displayed are averages of individual mouse weight changes from day 0 divided by weight on day 0. The plasmid harbored by the infecting strain is indicated in the legend. * are Welch t-test values ≤ 0.05 FDR controlled using the two-stage step up method of Benjamini, Krieger, and Yekutieli. (E, F) Fecal toxin B levels on days 1 and 4, determined by ELISA. Individual mice are displayed by a point, and a box and whisker plot shows the mean, quartiles, and extremes for cohorts with ≥3 toxin positive samples. * are significant FDR-controlled Kruskal-Wallis p-values < 0.05, ** < 0.005, FDR controlled using the two-stage step up method of Benjamini, Krieger, and Yekutieli.

While clindamycin-exposed mice challenged with either strain showed high levels of fecal toxin B at 24h after *C. difficile* challenge (Fig 5D), toxin levels in cefoperazone-exposed mice evolved more slowly, with peak levels occurring 4 days after exposure to ATCC 43255 pCD41 (Fig 5E). These findings demonstrate how increasing doses of cefoperazone progressively deplete the gut microbiota to enhance host susceptibility to *C. difficile*, particularly with strains harboring *mreE* variants that enable pathogen colonization and growth under the selective pressures provided by 3rd generation cephalosporins.

## Discussion

We used patient isolates from a *C. difficile* genomic surveillance program to discover *mreE* variants that increase resistance to ampicillin and to first and third-generation cephalosporins, including when transferred *in trans* to the susceptible strain CD630. Though some parent strains demonstrated reistance to cefepime, resistance was not transferrable *in trans* with the *mre* locus to strain CD630, suggesting that additional genomic factors may contribute to resistance to 4th generation cephalosporins. In addition, the *mreE* variants remained susceptible to 2nd generation cephlosporins and to the carbapenem meropenem.

Third-generation cephalosporins were the most commonly prescribed antibiotics in our patient cohort. Notably, the detection of cephalosporin-resistant *mreE* variants in *C. difficile* was associated with patient cephalosporin exposures 8-10 days prior to the detection of resistant strains, suggesting a critical window in which cephalosporin exposures increase patient risks for colonization with resistant strains and development of CDI. The broad distribution of *mreE* alleles among diverse genomic sequence types also suggests that antibiotic selective pressures may drive their evolution and dissemination, in both inpatient and outpatient settings (Fig. S4).

Introduction of the highly resistant *mre* locus from strain CD41 into the mouse-infective strain ATCC 43255 enabled symptomatic infection during cefoperazone administration at the selective dose of 500ug/mL, but not at the non-selective dose of 250ug/mL which limited microbiota depletion. The dose of 500ug/mL of cefoperazone in drinking water falls within range of doses used therapeutically in patients, and that can both ablate protective gut commensals and promote selection of antibiotic-resistant strains of *C. difficile* to colonize gut ecosystems.

The molecular mechanisms by which MreE mutations increase resistance to specific classes of beta-lactam antibiotics [7][8, 9] are as yet undefined and may include reduced binding affinity of affected antibiotic classes, in addition to interaction with other genomic loci that can produce higher levels of resistance, a finding suggested by the fact that transfer of the *mreE* alleles alone into susceptible strains did not elevate most cephalosporin MICs to the level of the original parent strain (Fig 2C). The gene-level variants also provide targets to aid in the detection of cephalosporin resistance, whether by strain genomic analyses or with targeted *mreE* probes to evaluate primary patient samples for resistant strains. Our identification of the window in which patient exposures to 3rd generation cephalosporins increased risks for *C. difficile* colonization and CDI also offers opportunities to inform programs in antimicrobial stewardship and for the monitoring of populations that are highly susceptible to CDI, to reduce risks.

In 1977, Bartlett, et al. identified toxigenic *C. difficile* as the driver of pseudomembranous colitis in clindamycin-exposed patients, by using patient isolates in clindamycin-exposed hamsters to capitulate disease [39]. In the deacdes since, strain phenotypic and genomic findings enhance our ability to discover how strain factors interact with antibiotic-depleted microbiota, and with host factors, to modulate *C. difficile’s* colonization and progression to disease [40]. Capacity to conduct comparable studies for other antibiotics, and more broadly for other pathogens, offers opportunities to mitiage patient risks for acquiring drug-resistant pathogens that increase risks for severe infections, particularly among vulnerable patient populations that experience high morbidity and mortality from such events.

## Data Availability

All data produced in the present work are contained in the manuscript

## Acknowledgements

We would like to thank Mike Feldgarden and Bill Klimke at NCBI for conversations about antimicrobial resistance that inspired these studies.

## Funding

These studies were funded by a Pilot and Feasibility Grant (Worley) from the Harvard Digestive Diseases Center P30 DK034854, R01 AI153605 (Bry) and the Hatch Family Foundation. The work of Jay Worley was supported by the National Center for Biotechnology Information of the National Library of Medicine (NLM), and the National Institute of Allergy and Infectious Diseases, National Institutes of Health.

---

**Fig S1.**
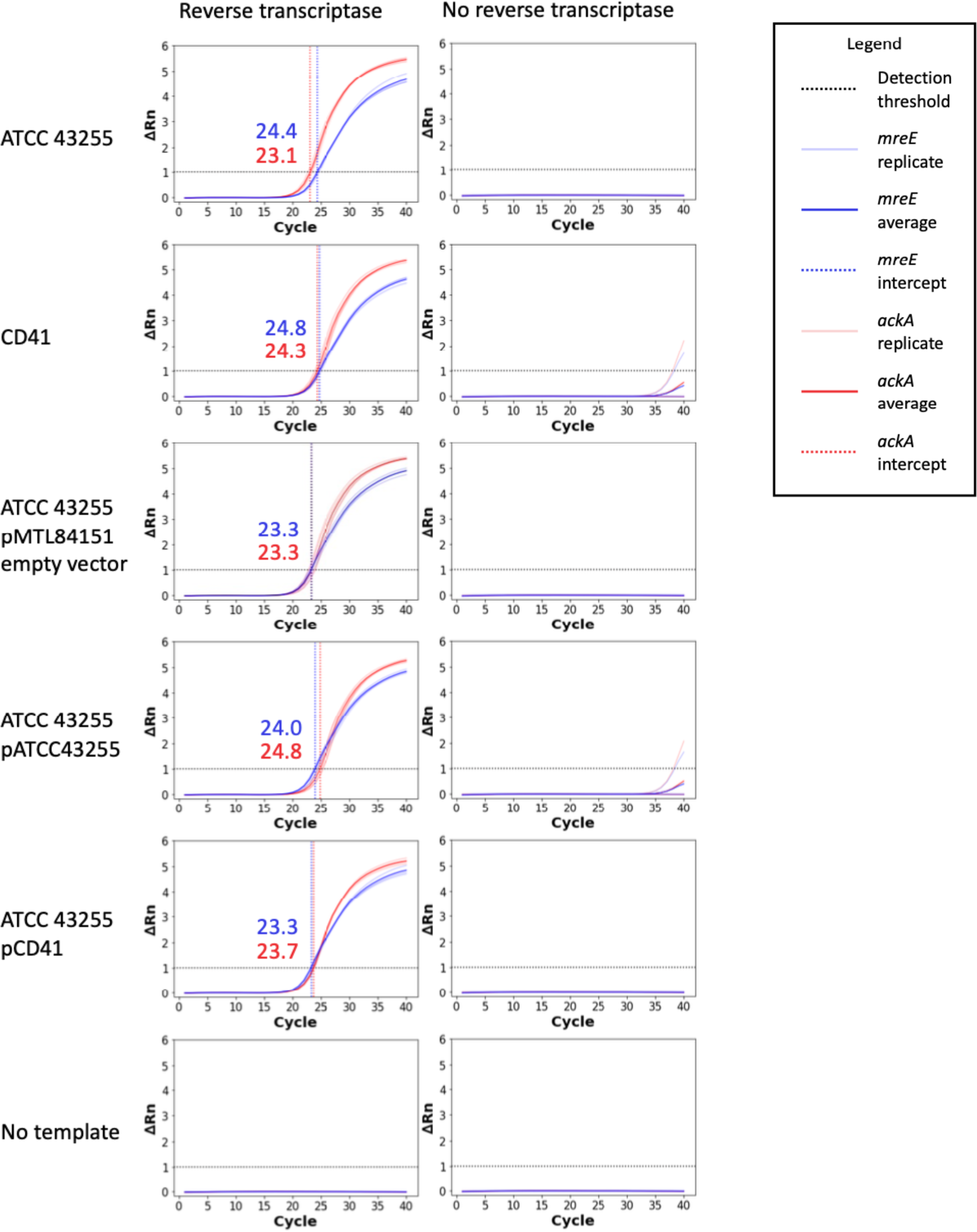
pMTL84151::*mre* does not overexpress *mreE.* The left column shows relative expression levels to the housekeeping gene *ackA* at mid-log phase using a common detection threshold at ΔRn 1 in a SYBR Green reverse-transcriptase PCR amplification assay. Right column is the same samples as the left column with no reverse transcriptase added. Samples crossing the detection threshold have the Ct value written in the graph in the corresponding color. The names pCD41 and pATCC43255 are pMTL84151 with an *mre* operon insert from the corresponding strain.

**Fig S2.**
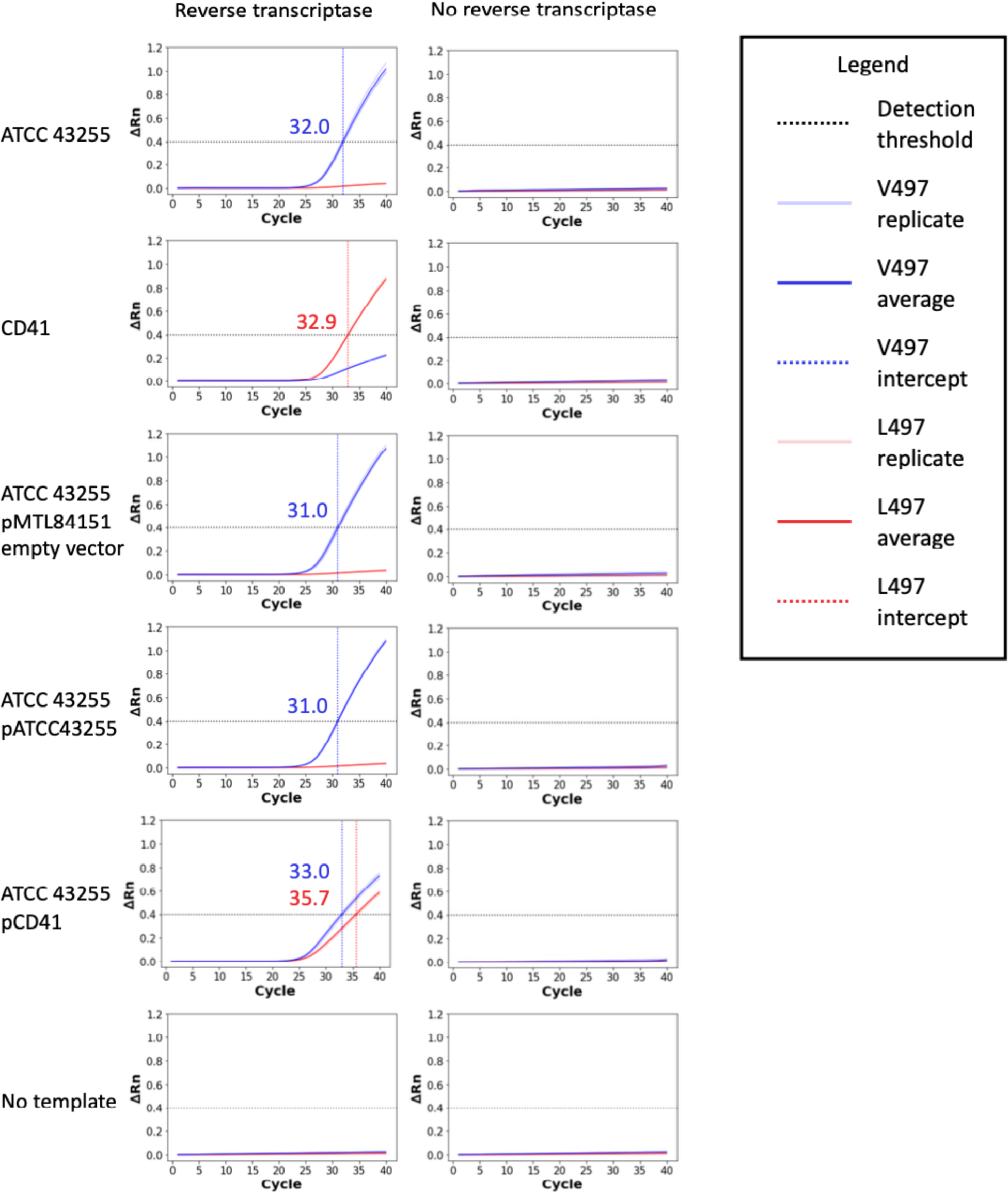
pMTL84151::*mre mreE* expression, across strains, is comparable to expression from the native chromosomal allele. The left column shows the relative expression levels at mid-log phase of the V497 and L497 encoded protein variants, representing the *mre* operon from ATCC 43255 and CD41, respectively. Expression was assayed using a probe-displacement PCR assay. Only ATCC 43255 pCD41 encodes both variants. The right column is the same samples as the left column with no reverse transcriptase added. Samples crossing the detection threshold have the Ct value written in the graph in the corresponding color.

**Figure S3.**
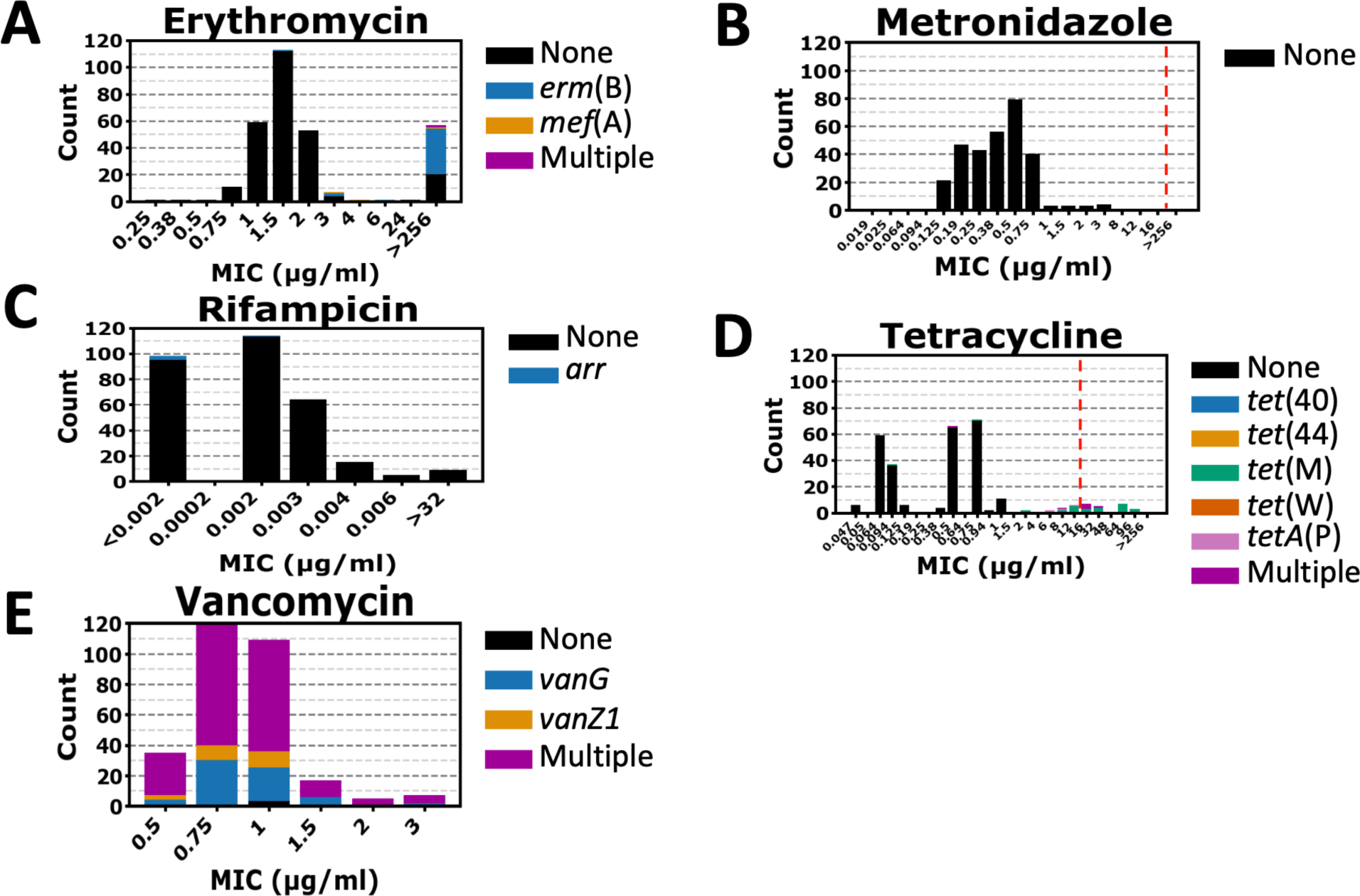
Additional antibiotic resistance tests and resistance gene correlations in clinical *C. difficile* isolates. The same isolates from Fig. 1A and B (n=306) assayed for antibiotic MIC and corresponding antibiotic resistance genes. (A) Erythromycin, (B) metronidazole, (C) rifampicin, (D) tetracycline, and (E) vancomycin. Corresponding antibiotic resistance gene presence is indicated by proportional coloring in each bar. If applicable, the CLSI breakpoint is indicated by a dashed red line.

**Figure S4.**
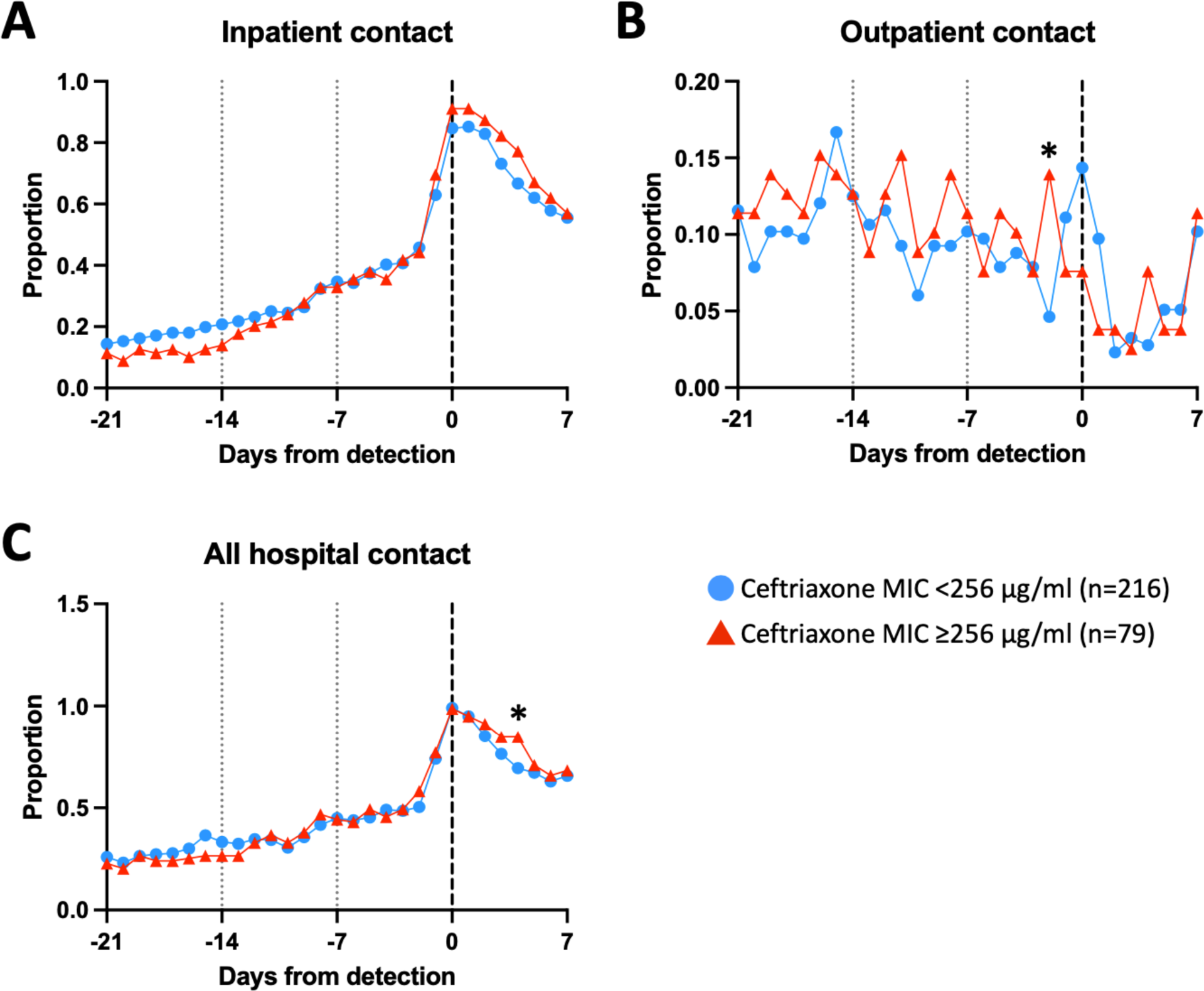
Strains with higher and lower ceftriaxone MICs have roughly equal hospital contact around the time of *C. difficile* detection. Patient cohort is the same as Fig. 1C and 1D (n=295). (A) Proportion of patients in each cohort receiving inpatient care on each day. (B) Proportion of patients receiving outpatient care on each day. (C) Proportion of patients receiving any hospital care on each day. * indicates a p-value < 0.05 by Fisher’s exact test for the population on the corresponding day.

**Figure S5.**
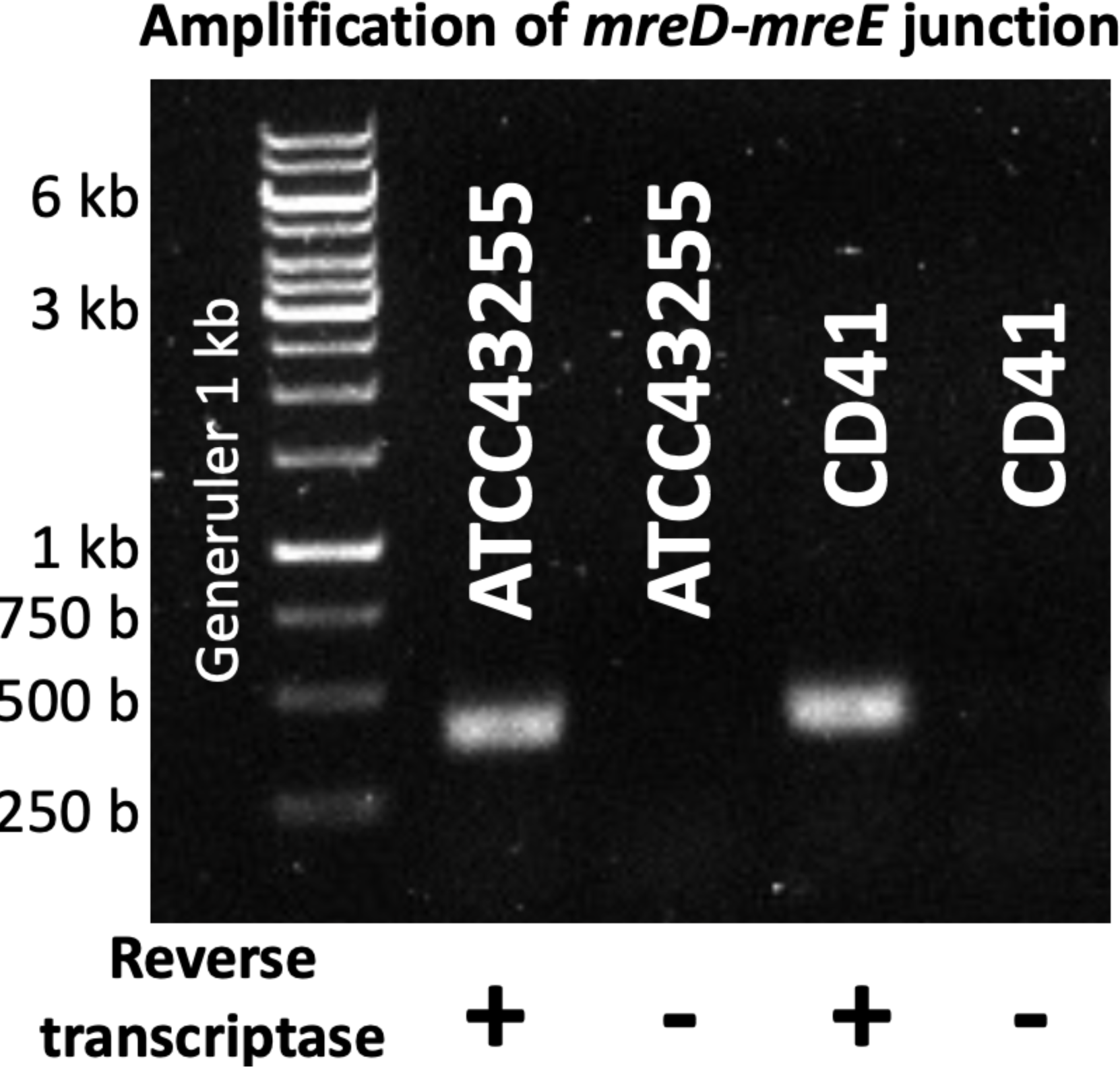
*mreE* is co-transcribed on a single mRNA molecule with *mreD*. PCR products using primers jnw-bwh-061 and jnw-bwh-062 amplifying a 432 bp amplicon including 244 bp of *mreD* and 177 bp of *mreE*, including the 11 bp intergenic region. Reverse transcriptase reactions used purified RNA from log phase cultures as template. Bands were only apparent when reverse transcriptase was added. Type strain ATCC 43255 and clinical strain CD41 are shown. Results demonstrate that the two open reading frames exist on a single mRNA molecule.

**Figure S6.**
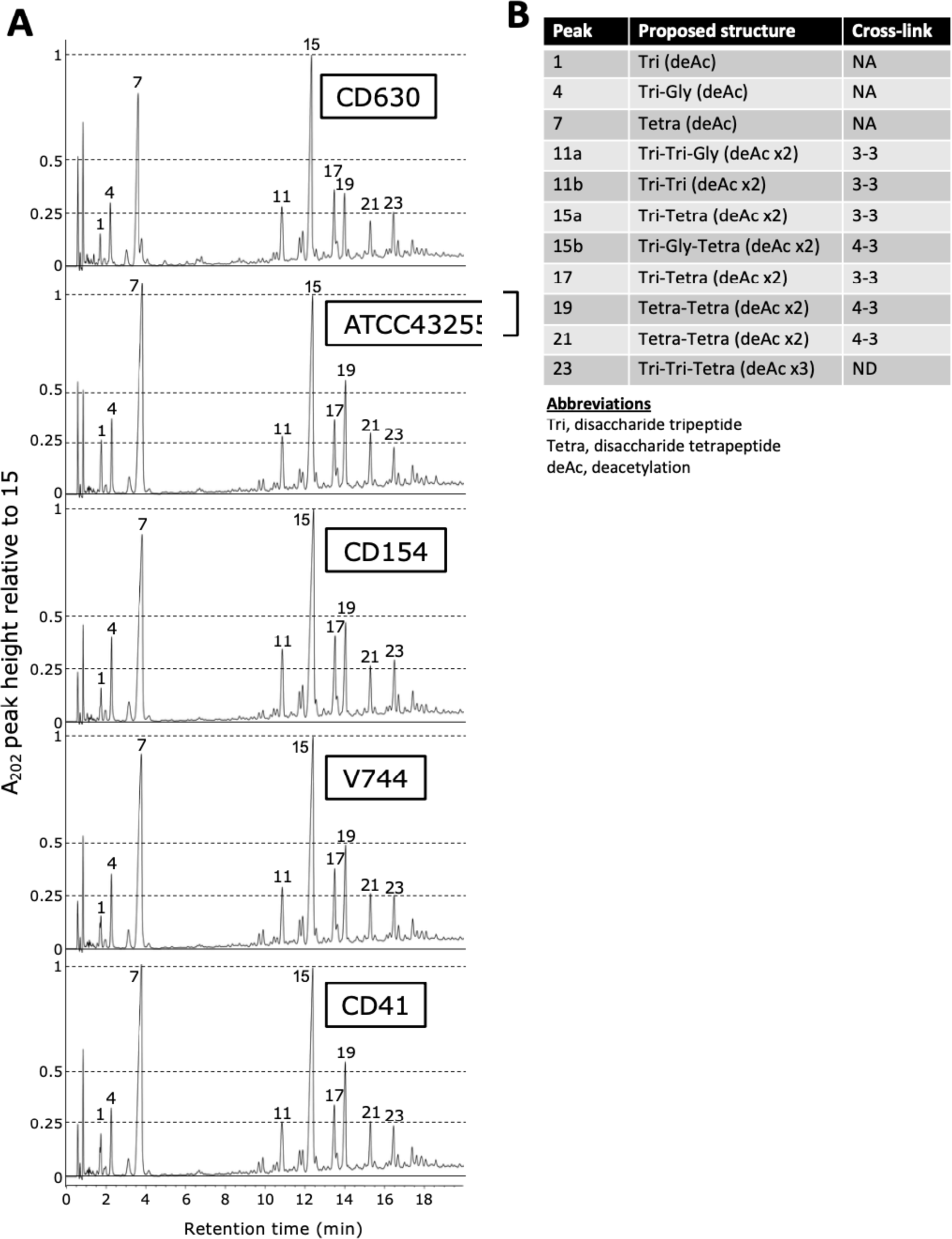
Peptidoglycan profiles in MreE variants strains. (A) Reverse-phase ultra-high-performance liquid chromatography (RP-UHPLC) chromatograms of muropeptides generated by mutanolysin hydrolysis *C. difficile* peptidoglycan for the given strain, showin in the box in the uppre right-hand corner, assayed by absorption at 202nm. Peak numbers correspond to those found in Peltier, et al., [21]. (B) Corresponding muropeptide structures. Peaks 11 and 15 contain two muropeptides, named “a” and “b”. Cross-link types are those connecting muropeptide dimers.

**Fig. S7.**
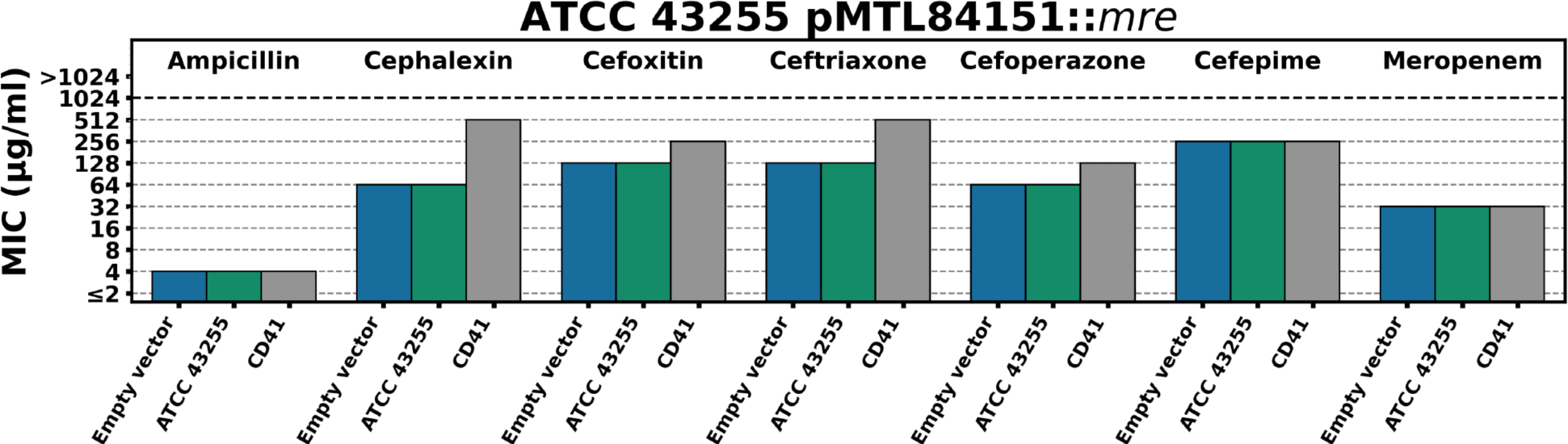
pMTL84151::*mre*_CD41_ increases cephalosporin resistance in the mouse infective strain ATCC 43255. MICs were determined, as described, by broth microdilution assay.

**Figure S8.**
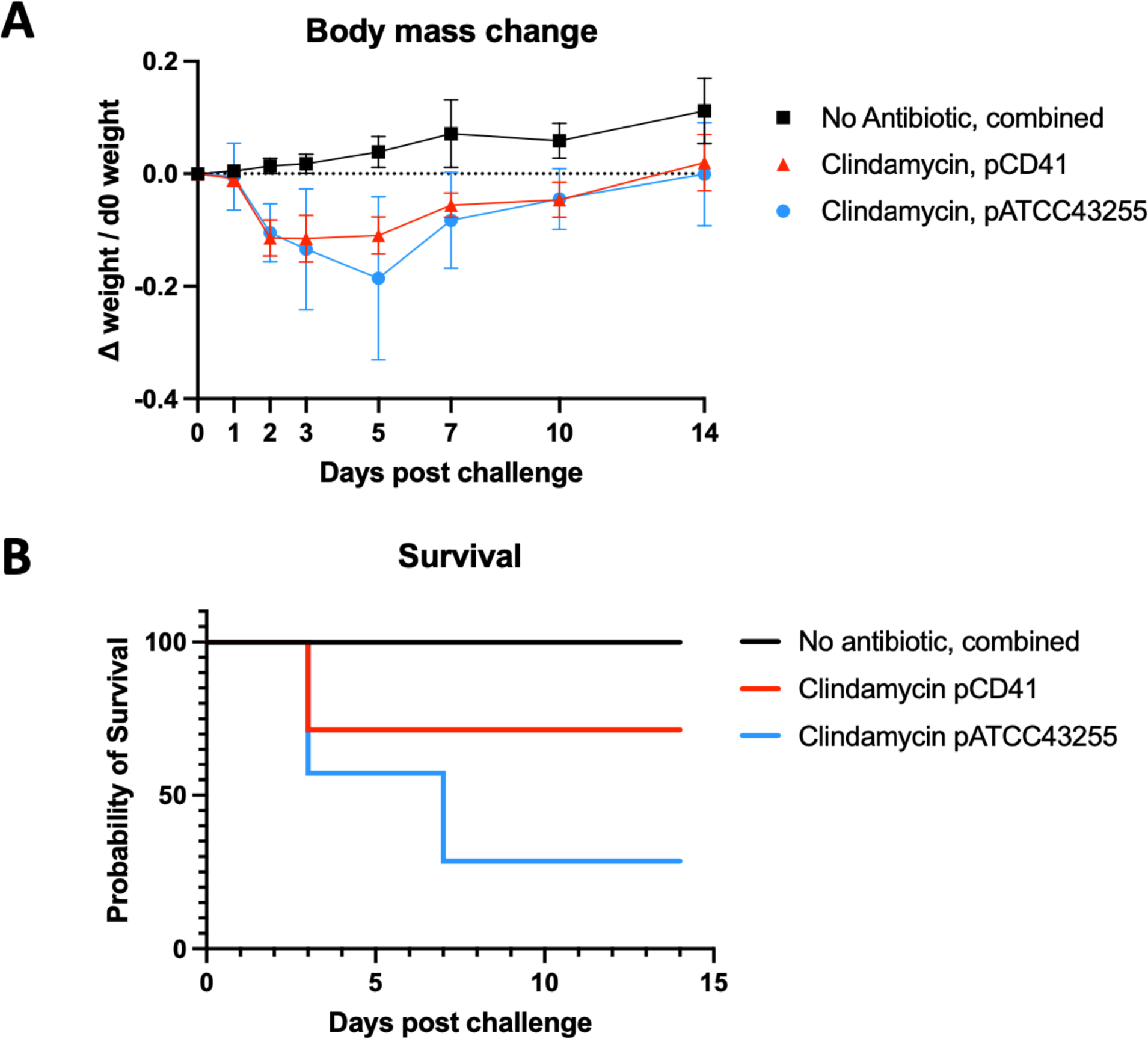
*C. difficile* ATCC 43255 isolates carrying *mre* operons on pMTL84151 are pathogenic in a standard mouse model of antibiotic-induced CDI and do not cause weight loss in mice untreated by antibiotics. (A) Average change in mouse weights from day 0 normalized to mouse weight on day 0. Error bars are one standard deviation. The “No Antibiotic” cohort contained both ATCC 43255 pCD41 (n=3 mice) and ATCC 43255 pATCC43255 (n=3 mice) infected mice. Both cohorts with clindamycin treatment had 7 mice. (B) Survival curve of the number of surviving mice in each cohort on the days in Figure S7A.

**Table S1.**
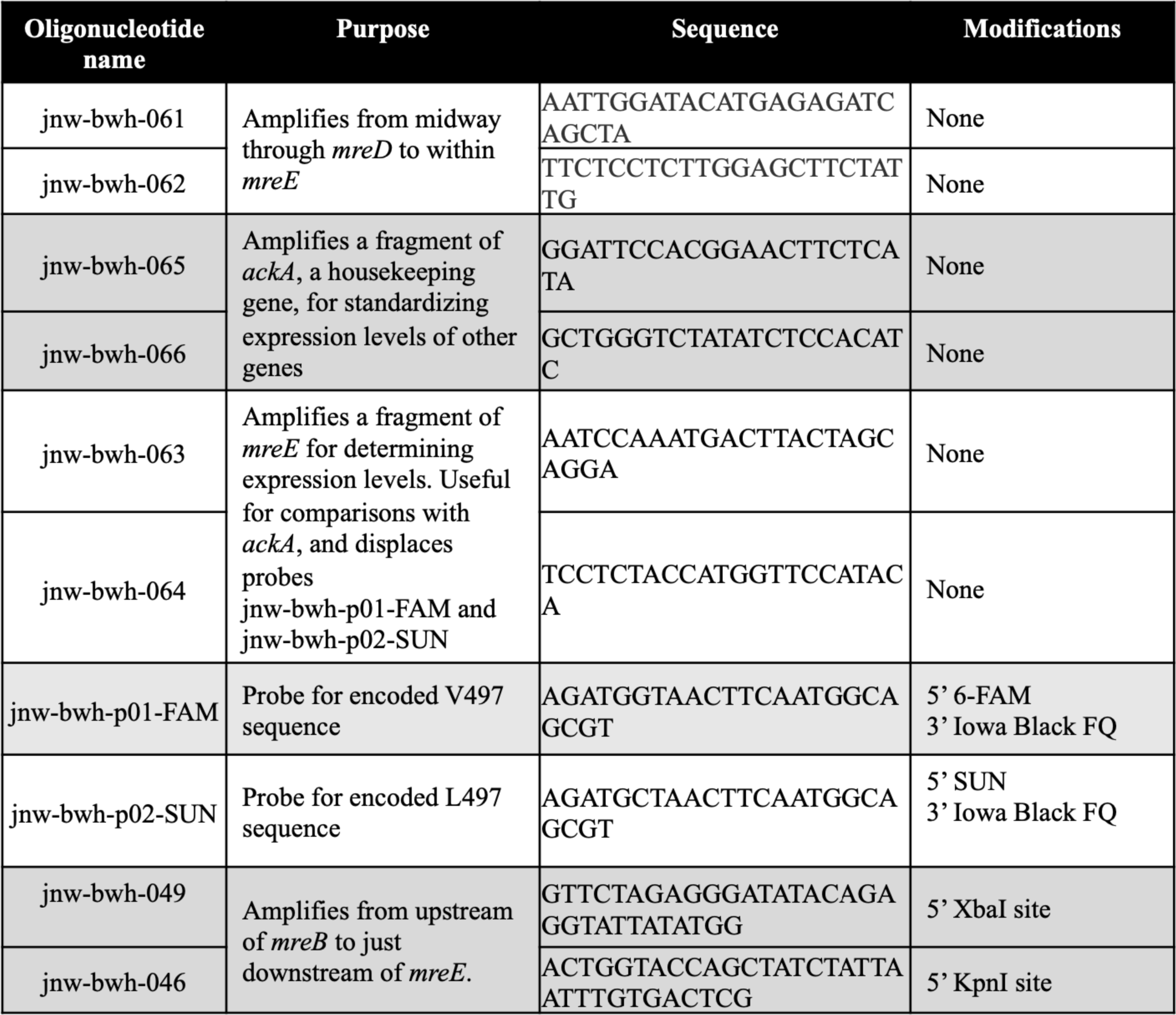
Probes and primers used in this study. Vertically adjacent rows show the forward and reverse amplification pairs.

**Table S2.** Protein sequence clades significantly associated with ceftriaxone resistance. Odds ratios and p values for cell wall synthesis loci associated with phenotypic ceftriaxone resistance. Loci associated with an odds ratio of five or higher are shown. * “Clade A” MreE variants; ** “Sub-clade A1” MreE variants.

| Locus | Protein Name | Odds Ratio | p value | In Clade,<br>Resistant | In Clade,<br>Susceptible | Out of Clade,<br>Resistant | Out of Clade,<br>Susceptible | BH Critical<br>Value | Adjusted p<br>value |
| --- | --- | --- | --- | --- | --- | --- | --- | --- | --- |
| 1496.5150.peg.2682 | MreE* | 113.7096774 | 1.14E-30 | 47 | 3 | 31 | 225 | 6.71E-05 | 1.70E-28 |
| 1496.5150.peg.2682 | MreE** | ≥107.5 | 5.22E-17 | 25 | 0 | 53 | 228 | 0.000134228 | 3.89E-15 |
| 1496.5150.peg.2043 | VanG | 39.42307692 | 2.82E-11 | 20 | 2 | 52 | 205 | 0.000201342 | 1.40E-09 |
| 1496.5150.peg.2041 | VanT | 24.37735849 | 7.38E-10 | 19 | 3 | 53 | 204 | 0.00033557 | 2.20E-08 |
| 1496.5150.peg.2043 | VanG | 10.98901099 | 3.70E-08 | 20 | 7 | 52 | 200 | 0.000469799 | 7.87E-07 |
| 1496.5150.peg.2045 | VanR | 9.142857143 | 2.07E-10 | 30 | 15 | 42 | 192 | 0.000268456 | 7.72E-09 |
| 1496.5150.peg.440 | Diaminopropionate ammonia-lyase | 6.60130719 | 1.33E-07 | 25 | 15 | 51 | 202 | 0.000536913 | 2.48E-06 |
| 1496.5150.peg.842 | NagX | 5.96719457 | 3.16E-07 | 25 | 17 | 52 | 211 | 0.000604027 | 5.23E-06 |
| 1496.5150.peg.442 | N-acyl-D-amino-acid deacylase | 5.854605993 | 4.03E-07 | 25 | 17 | 53 | 211 | 0.000671141 | 6.00E-06 |
| 1496.5150.peg.770 | DltB | 5.854605993 | 4.03E-07 | 25 | 17 | 53 | 211 | 0.000738255 | 5.46E-06 |
| 1496.5150.peg.909 | PdaA | 5.854605993 | 4.03E-07 | 25 | 17 | 53 | 211 | 0.000805369 | 5.00E-06 |
| 1496.5150.peg.2901 | Metallo-beta-lactamase<br>superfamily protein | 5.854605993 | 4.03E-07 | 25 | 17 | 53 | 211 | 0.000872483 | 4.62E-06 |

## Notes

### Competing Interest Statement

The authors have declared no competing interest.

### Author Declarations

The Mass-General-Brigham (MGB) IRB gave ethical approval for this work.

